# Cilengitide limits the progression of both syndromic and sporadic thoracic aortic aneurysms by targeting alpha V integrins

**DOI:** 10.1101/2025.03.21.25324435

**Authors:** Sara Rega, Silvia Bouhuis, Nadia Fanotti, Manuel Casaburo, Simone Vodret, Francesco Riccitelli, Federico Celotto, Luca Lambroia, Montserrat Climent, Leonardo Elia, Martina Vescio, Linda Pattini, Rosaria Santoro, Veronika Myasoedova, Paolo Poggio, Giorgia Bonalumi, Nathasha Samali Udugampolage, Jacopo Taurino, Alessandro Pini, Serena Zacchigna, Giulio Pompilio, Gianluca L. Perrucci

## Abstract

Thoracic aortic aneurysm is a life-threatening condition due to either genetic syndromes (*e.g.*, Marfan syndrome) or cardiovascular risk factors (*e.g.*, hypertension, aging and smoking), which favour the onset of sporadic thoracic aneurysms. Activation of the transforming growth factor-β pathway and dysregulation of mechanotransduction signals in vascular smooth muscle cells play a key role in the development of both syndromic and sporadic forms of thoracic aortic aneurysm. The precise molecular mechanisms underlying thoracic aortic aneurysm onset and progression are still unresolved and available therapies merely rely on surgical intervention.

Integrins containing the αV subunit are central to both transforming growth factor-β (TGF-β) and mechanotransduction signalling pathways, leading to pro-fibrotic molecular events. Here we investigate the role of αV integrins in the development of both syndromic and sporadic thoracic aortic aneurysms and the therapeutic potential of two αV integrin inhibitors (Cilengitide and GLPG0187). We observed that αV integrins are more expressed in both types of human thoracic aortic aneurysms and that integrin inhibition limits TGF-β activation and mechanotransduction-related pro-fibrotic pathways in patient-derived vascular smooth muscle cells. *In vivo* experiments revealed that Cilengitide is the most effective αV integrin inhibitor in limiting the dilation of the aortic bulb in murine models of both syndromic and sporadic forms of thoracic aortic aneurysms. These findings set the αV integrin inhibitor Cilengitide as a promising drug for the treatment of thoracic aortic aneurysms.

## 1. Introduction

Thoracic aortic aneurysm (TAA) consists in a weakened and dilated portion of the thoracic aorta, often resulting in more than 50% increase in the aortic diameter and in life-threatening complications, such as aortic dissection or vessel rupture [1]. Depending on their aetiology, TAA are classified in three main forms: (*i)* sporadic TAA (sTAA), often caused by the interplay between common cardiovascular risk factors (e.g. hypertension, aging and smoking) and *de novo* genetic mutations, (*ii)* familial TAA, which is a genetically determined, but not associated with other pathological manifestations and (*iii)* syndromic TAA, due to rare, genetic mutations leading to multi-organ syndromes, such as Ehlers-Danlos, Loeys-Dietz and Marfan syndrome (MFS). The global incidence of TAA is 5-10 *per* 100,000 persons years, accounting for 150,000-200,000 deaths *per* year [2]. Despite this burden, available therapies are merely palliative and surgical replacement of the thoracic aorta with a prosthetic graft remains the only effective option [3, 4]. Therefore, the characterization of key molecular mechanisms leading to aortic dilation in both conditions is necessary to identify relevant targets and develop innovative and etiological therapies [5].

Over the past 25 years, several studies have started shedding light on the molecular mechanisms that underlie TAA development, which include transforming growth factor-beta (TGF-β) signalling [6–8] and mechanotransduction response of aortic vascular smooth muscle cells (VSMC) to extracellular matrix (ECM) alterations [9, 10].

A key role in mechanotransduction is played by integrins, heterodimeric transmembrane receptors, composed by “α” and “β” subunits, which act as molecular “bridges” between ECM proteins and the intracellular space [9, 11]. Among integrin family members, the αV subunit is of particular interest in TAA as i) it releases active TGF-β from the ECM [12] and ii) it has high affinity for specific thoracic aortic ECM proteins, containing the Arginine-Glycine-Aspartate (RGD) sequence, such as fibronectin, vitronectin and fibrillin-1 [13]. Moreover, αV integrins are known to modulate multiple mechanosensing pathways (*e.g.,* Rho GTPase signalling) [14–16]. Both TGF-β and mechanotransduction signalling pathway are key drivers of pro-fibrotic response in multiple cell types, tissues and organs [17, 18]. We and others have previously reported that pharmacological inhibition of αV integrins in rat cardiac fibroblasts prevents their differentiation into myofibroblasts and production of pro-fibrotic molecules [19] and attenuates fibrosis in a mouse model of cardiac fibrosis and systemic sclerosis [20, 21]. Enhanced expression of β3 integrin was reported in a murine model of MFS (*Fbn1*^C1039G/+^) [22], and αV integrin overexpression was detected in human iPSC-derived MFS-VSMC [22]. Additionally, systemic αV integrin inhibition, by GLPG0187, reduced FAK-Akt-mTORC1 signalling, mitigated VSMC pro-fibrotic phenotypic switch and limited aneurysm formation in *Fbn1*^C1039G/+^ mice [23]. Altogether these data confirm an important involvement of integrins, and in particular of αV integrin, in TAA. However, the role of integrins in human primary VSMC, and specifically in the context of sTAA, has not yet been reported. Here, we wish to address this gap, by analysing human thoracic aortic specimens from healthy control (HC), MFS-TAA and sTAA patients assessing the role of two integrin inhibitors in both MFS and sTAA murine models.

## 2. Materials and Methods

### 2.1. Data Availability

Data of this study are available from the corresponding author upon reasonable request on ZENODO (doi: 10.5281/zenodo.15064795).

### 2.2. Patient Enrolment

Both dilated and non-dilated portions of thoracic aortic aneurysms were collected from non-syndromic sporadic (sTAA) and syndromic (MFS) patients underwent surgical aortic replacement. Samples of sTAA patients were collected in the surgery room of Centro Cardiologico Monzino IRCCS, while MFS patients’ specimens from surgery room of ASST FBF-SACCO. All patients signed and gave written informed consent for tissue collection for research purposes and the procedures were approved by Ethics Committee either of Centro Cardiologico Monzino IRCCS (RE3001-CCM 1462) and ASST FBF-Sacco (Prot. N. 39138/2016, Milan, Italy). Sensitive data were handled in accordance with the Helsinki Declaration. The healthy aortic samples were provided by Tebubio (Tebubio, Milan, Italy). Fresh biopsies were divided into fragments dedicated to histology, cells isolation and molecular studies. Patients’ generalities have been reported in **Supplementary Table S1**. Patients undergoing aortic repair, of either sex, aged 18 years or older, who have read and signed the informed consent have been included in the study. On the contrary, patients with previous aortic and/or mitral valve surgery, early menopause and/or osteoporosis, rheumatic heart disease, active malignancies, with acute and chronic inflammatory states, chronic liver disease, chronic renal failure (creatinine > 1.5 mg/dl), and thyroid-related diseases have been not enrolled in the study.

### 2.3. Spatial transcriptomics

Aortic fragments dedicated to histology analyses were fixed with Carnoy’s solution (60% ethanol, 30% chloroform and 10% glacial acetic acid) and paraffin-embedded (CFPE). Then, for spatial transcriptomics experiments, 3-μm-thick sections were cut from each aortic wall sample. After deparaffinization and rehydration, sections were stained with haematoxylin for 15 minutes and eosin for 7 minutes (Bio-Optica, Milan, Italy). RNA was extracted using the RNeasy FFPE Kit (Roche, Basel, Switzerland), following the manufacturer’s instructions, and DV200 values were evaluated on a TapeStation (Agilent, Santa Clara, CA, USA) by using the High Sensitivity RNA ScreenTape Kit (Agilent). Only FFPE vessel blocks with a DV200 > 50 were included in the analysis. CFPE aneurysm samples were processed according to the manufacturer’s guidelines. H&E images were prepared following the same protocol, and brightfield histologic images were acquired using the AxioScan.Z1 (ZEISS). Libraries were constructed using the Visium Spatial for CFPE Gene Expression Kit (10X Genomics, cat. no. 1000336) and sequenced on a NextSeq2000 (Illumina, San Diego, CA, USA) with a minimum sequencing depth of 50,000 read pairs *per* spatial spot. Sequencing was conducted as recommended (read 1: 28 cycles; i7 index read: 10 cycles; i5 index read: 10 cycles; read 2: 50 cycles), yielding 200–230 million reads *per* section. Reads were mapped to the GRCh38 reference genome and aligned using Spaceranger v2.0.0 (10X Genomics). Downstream analysis was performed in R v4.2.3 using Seurat v5.1.0. Spots capturing fewer than 200 features were filtered, and the data were normalized using the SCTransform method. Clustering was performed with the Seurat functions FindNeighbors (top 30 dimensions) and FindClusters (resolution 0.3). Scores for pathway of interest were assigned to spots based on literature-derived gene sets using the AddModuleScore function. Spot composition inference was conducted by integrating single-cell RNA data from Tabula Sapiens [24] with the NNLS method in semla. Aggregated pseudobulk transcriptomic data were used to identify differentially expressed genes with EdgeR v3.42.4, using the glmLRT test.

### 2.4. Immunostaining and Histological Assays

5-μm-thick CFPE sections were rehydrated, incubated with primary antibodies against αV integrin (AbCam, Boston, MA, USA), and COL1A1 (Cell Signaling Technology, Danvers, MA, USA) over-night (O/N) at 4°C, and secondary antibodies conjugated with AlexaFluor fluorophores for 1 h at room temperature (RT). Nuclear staining was performed with DRAQ5 or Hoechst 33342 Buffer (Life Technologies, Carlsbad, CA, USA). Slides were viewed with ApoTome2 microscope equipped with AxioCam camera (Carl Zeiss, Oberkochen, Germany). Densitometric analyses measurements were performed with ZEN Blue software (Carl Zeiss, Oberkochen, Germany).

### 2.4. Aortic VSMC Isolation

Samples obtained from patients underwent thoracic aortic surgical substitution were abundantly washed with Phosphate Buffer Saline (PBS) for blood removal. *Media* and *adventitia tunicae* were physically separated, then *tunica media* was minced and digested O/N at 37°C in a solution of 2 mg/mL collagenase type II (Worthington Biochemical Corporation, Lakewood, NJ, USA). The result of tissue digestion was filtered the day after with 100 μm cell strainer, pelleted and plated in Smooth Muscle Cell Growth Medium-2 Singlequots (SmGM-2, Lonza, Basel, Switzerland). To improve cell growth, the day after isolation the medium was changed to remove residual erythrocytes and ECM debris.

### 2.5. ImageStreamX Imaging Flow Cytometry

For αV, β5, αVβ3, α5, and β1 integrins cellular localization experiments, cells were acquired by the ImageStreamX imaging cytometer (Amnis Corporation, Seattle, WA, USA), with 40X of magnification and low flow rate/high sensitivity using the INSPIRE software. Cells were gently detached from Petri dishes by using the TrypLE Select solution (ThermoFisher Scientific, Paislay, Scotland, UK). Afterward, cells were pelleted, resuspended in FACS buffer (1X PBS, 0.1% Bovine Serum Albumin (BSA), and 5 mM EDTA) and incubated with conjugated primary antibodies for 10 minutes at RT. Data from 10,000 events *per* sample were collected and the percentage of the positive elements was calculated using the IDEA software (Amnis Corporation, Seattle, WA, USA). Primary antibodies against αV-FITC, β5-PE, αVβ3-APC, α5-APC and β1-PE (Miltenyi Biotec, Bergisch Gladbach, Germany) were used.

### 2.6. Western Blot

Both aortic tissues and cells were lysed in cell lysis buffer (Cell Signaling Technology, Danvers, MA, USA) supplemented with protease and phosphatase inhibitor cocktails (Sigma-Aldrich, Saint Louis, MO, USA). Cells were previously treated with vehicle (complete medium), 5 ng/ml of TGF-β1 (PeproTech, London, UK), 0.5 µM Cilengitide (MedChemExpress, Monmouth Junction, NJ, USA), 0.5 ng/ml GLPG0187 (Cayman Chemicals, MI, USA). Total protein extracts were subjected to SDS-PAGE and transferred onto a nitrocellulose membrane. The membranes were blocked for 1 h at RT in 5% non-fat dry milk in Wash Buffer (1X Tris Buffer Sulfate, 0.1% Tween 20) and then incubated O/N at 4°C with the appropriate primary antibody. Primary antibodies used were specific for: αV integrin (AbCam, Boston, MA, USA), COL1A1, phospho-SMAD2/3, SMAD2/3, phospho-ERK1/2, ERK1/2, RhoA, ROCK1, αSMA (Cell Signaling Technology, Danvers, MA, USA), and GAPDH (Santa Cruz Biotechnology, Dallas, TX, USA). The membranes were incubated with peroxidase-conjugated secondary antibodies (GE Healthcare, Chicago, IL, USA) for 1 h. Signals were visualized using the enhanced chemiluminescence Western blotting detection system (Bio-Rad Laboratories, Hercules, CA, USA). Images were acquired with Chemidoc system and densitometric analysis of membranes was performed using the ImageLab Software (Bio-Rad Laboratories, Hercules, CA, USA). GAPDH has been used as protein loading control. To analyse functional RhoA levels, the Rho-GTPase Antibody Sampler Kit (Cell Signaling Technology Danvers, MA, USA) was used following the manufacturer’s protocol.

### 2.7. Immunofluorescence

Cells were seeded on round glasses, coated with recombinant human vitronectin protein (rh-VTN, Gibco), and treated as previously described for 48 hours. Then, glasses were washed in PBS solution, and fixed with 4% paraformaldehyde (PFA, Sigma-Aldrich, St. Louis, MO, USA) for 15 min. Cells were permeabilized with 1X PBS supplemented with 5% BSA and 0.1% Triton X-100, and then incubated O/N at 4°C with the primary antibody raised against αV integrin (AbCam, Boston, MA, USA), and COL1A1 (Cell Signaling Technology, Danvers, MA, USA). The goat anti-mouse IgG (H+L) secondary antibody conjugated with AlexaFluor594 (ThermoFisher Scientific, Paislay, Scotland, UK), and goat anti-rabbit IgG (H+L) secondary antibody conjugated with AlexaFluor488 (ThermoFisher Scientific, Paislay, Scotland, UK) were incubated for 1h at RT. Nuclei were stained with Hoechst 33342 Buffer (Life Technologies, Carlsbad, CA, USA). Glasses were mounted on a slide by DAKO Mounting Medium for immunofluorescence (Sigma-Aldrich, Saint Louis, MO, USA). Slides were viewed with ApoTome2 microscope equipped with AxioCam camera and analysed with ZEN Blue sotfware (Carl Zeiss, Oberkochen, Germany).

### 2.8. Animal models

Animal protocols were approved by the Italian Ministry of Health with the authorization n. 4/2022-PR. We adopted two different animal protocols, depending on the TAA model (*i.e.,* syndromic or non-syndromic sporadic TAA). Fbn1^C1039G/+^ mice were used to model MFS-TAA [6] and were purchased from the Jackson Laboratory (Bar Harbor, ME, USA), supplied by Charles River Laboratories Italia S.r.l. (Lecco, Italy). The sTAA model has been obtained by administering in drinking water the lysyl oxidase inhibitor β-aminopropionitrile (BAPN, 1 g/kg/day added to 5 ml H_2_O/mouse/day for 21 days) [25, 26]. Both TAA models and littermate control (C57BL/6J) mice were treated three times a week, intraperitoneally, with either Cilengitide 90 mg/kg/day (MedChemExpress, Monmouth Junction, NJ, USA), GLPG0187 100 mg/kg/day (Cayman Chemicals, Michigan, MI, USA) or vehicle (1% DMSO in PBS). The number of animals was calculated through G*Power (v. 3.1.9.2) software. Furthermore, a correction factor was added for each transgenic murine line, to overcome the animal loss due to non-specific mortality.

Animals were monitored with color-doppler 2D-echocardiography (Vevo3100, VisualSonics, Toronto, Canada, USA) equipped with a linear matrix transducer of 40 MHz (550D model), every 4 weeks (MFS mice) or weekly (sTAA mice), to analyse aortic and cardiac function. Animals were euthanized by anesthesia with Ketamine 100 mg/kg at 16 weeks of age (MFS mice) or at 9 weeks of age (sTAA), and their organs were collected to perform morphometric, histologic, molecular and biochemical analyses. For aortic root/ascending aorta samples, the aortic root was completely dissected from the left ventricle, cutting below the aortic valve and harvested considering the region until the brachiocephalic artery.

### 2.9. Aortic 2D-echocardiography

Transthoracic echocardiography was performed on mice sedated with 2% inhaled isoflurane (2-chloro-2-(difluoromethoxy)-1, 1, 1-trifluoroethane) (Baxter Healthcare Corporation, Deerfield, IL). Images of the aortic root and mid ascending aorta (halfway between the sinotubular junction to the brachiocephalic artery) were acquired in parasternal long-axis with an MS700 70 MHz MicroScan transducer on the Vevo 2100 system (VisualSonics, Toronto, Canada). The most dilated portion of the aortic root sinuses and ascending aorta were measured during end-diastole. The aortic measurement was performed by two blinded investigators in triplicate.

### 2.10. Masson’s trichrome staining

5-μm-thick sections of CFPE were cut from each sample. Sections were rehydrated, and Masson’s trichrome staining kit (Bio-Optica, Milan, Italy) was performed, following manufacturer’s protocol. Slides were viewed with AxioImager microscope (Carl Zeiss, Oberkochen, Germany). Densitometric analyses measurements were performed with ZEN Blue software (Carl Zeiss, Oberkochen, Germany), by two blinded readers.

### 2.11. Statistical Analyses

Quantitative results are expressed as mean±SD. Statistical significance was evaluated with GraphPad Prism 10. The comparisons between two groups were analysed by Students-*t* test, while more than two variables were analysed by one-way ANOVA with Tuckey’s *post-hoc* test or two-way ANOVA with Bonferroni’s *post-hoc* test, as appropriate. A value of *p*<0.05 was deemed statistically significant.

## 3. Results

### 3.1 αV integrin, ECM proteins and pro-fibrotic mediators are highly expressed in the thoracic aorta of patients with both MFS and sporadic TAA

We performed spatial transcriptomics on samples of thoracic aortas obtained from HC and patients affected by either MFS-TAA or sTAA, whose features are reported in **Supplementary Table S1**.

Data were processed in R, using the Seurat workflow [27]. Spots capturing fewer than 200 genes were excluded from the analysis. Data were integrated and subjected to clustering to identify groups of spots with similar transcriptional profiles. This approach allowed segmentation of tissue sections into 11 spot clusters, with shared transcriptomic profiles across patients, as shown in **Figure 1A**. Specifically, clusters 0, 1, 5, 7, and 9 mapped to the medial layer, cluster 3 to the *adventitia* and clusters 2, 4, 6, 8, and 10 to the *intima*. To disentangle the cellular composition of the analysed clusters, we interrogated the “vasculature” subset in the single-cell RNA *Tabula Sapiens* dataset [24], using the NNLS method implemented in the R package semla [28]. The average percentage of the complexity of cell types within each cluster is shown in **Figure 1B**. Next, we evaluated pathways known to be implied in TAA, including integrin signalling, ECM remodelling, TGF-β signalling, and mechanotransduction (**Figures 1C, D**). Genes involved in these pathways were significantly more expressed in sections from TAA patients, in both dilated and non-dilated regions, than in HC. Notably, ECM-related pathways were predominantly active in the *intima*, while integrin and TGF-β signalling were more prominent in the medial layer. Finally, differential gene expression analysis was performed by aggregating spot data into pseudo-bulk samples and comparing groups using EdgeR. This analysis confirmed the upregulation of integrin genes, such as *ITGAV*, *ITGB1*, *ITGB3*, *ITGB5* (**Figures 1E, F**; **Table 1**) and collagen remodelling genes (**Supplementary Figure S1**; **Table S2**) in both TAA patients *vs* HC.

**Figure 1.**
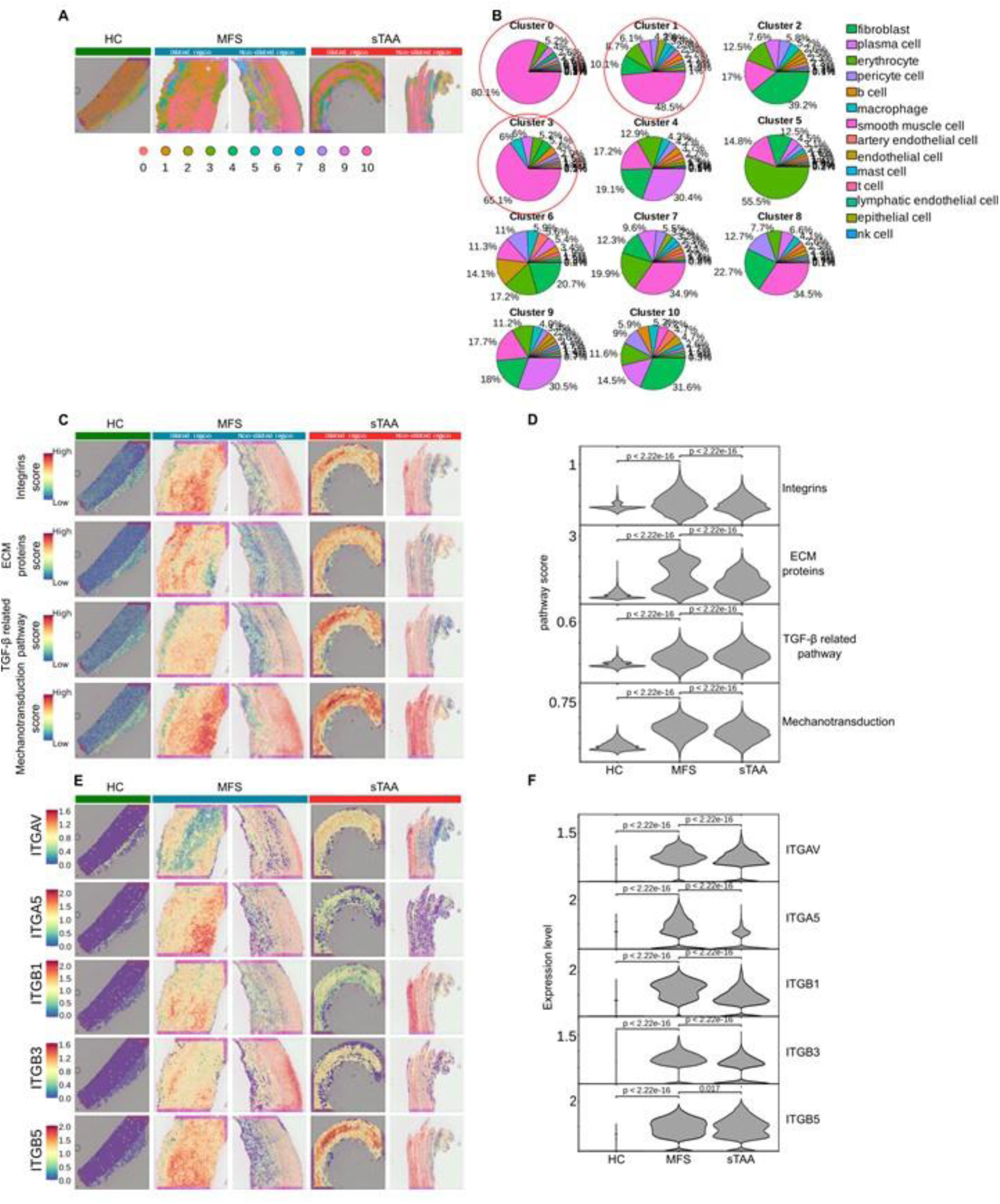
Specific gene pathways are upregulated in MFS-TAA and sTAA than in HC patients. (**A**) Images of spatial transcriptomic analysis on thoracic aortas from HC subjects and patients with MFS or sTAA, both in dilated and non-dilated aneurysmal regions. Images show the spatial distribution of 11 cluster of spots that share a similar transcriptional profile, reflecting a peculiar cell composition. (**B**) Pie charts showing the various cell types that are present in the 11 transcriptomic clusters, deconvoluted using single-cell RNA sequencing data from the *tabula sapiens* vascular dataset (https://tabula-sapiens-portal.ds.czbiohub.org). Clusters 0, 1 and 3 (circled) contain the highest number of VSMC and are mainly located in the tunica media in all aortic samples. (**C**) Images of spatial transcriptomic analysis showing transcript number density for the indicated pathways (blue: low number, red: high number). (**D**) Violin plots representing the quantification of the gene expression levels for the indicated pathways in the different samples. (**E**) Images of spatial transcriptomic analysis showing expression level of five genes encoding integrins (blue: low expression, red: high expression). (**F**) Violin plots representing the quantification of the expression levels for the integrin genes in the different samples.

**Table 1.** DEGs related to integrins obtained by spatial transcriptomics analysis data.

| DEGs (MFS vs HC) |  |  |  |  |  |
| --- | --- | --- | --- | --- | --- |
| Gene | logFC | logCPM | LR | PValue | FDR |
| *ITGAV* | 2,035966 | 7,601496373 | 12,17131281 | 0,000485301 | 0,014338611 |
| *ITGB1* | 2,666992 | 9,821758609 | 15,68732392 | 7,47233E-05 | 0,005731791 |
| *ITGB3* | 1,947082 | 7,009591047 | 16,0212606 | 6,26352E-05 | 0,005301094 |
| *ITGB5* | 2,007023 | 9,570096349 | 8,018246548 | 0,004630838 | 0,040669925 |
| DEGs (sTAA vs HC) |  |  |  |  |  |
| Gene | logFC | logCPM | LR | PValue | FDR |
| *ITGAV* | 2,122407 | 7,601496373 | 13,54647816 | 0,000232728 | 0,005696867 |
| *ITGB1* | 2,397938 | 9,821758609 | 13,50296007 | 0,000238187 | 0,005735837 |
| *ITGB3* | 1,60353 | 7,009591047 | 11,63231129 | 0,00064816 | 0,00971748 |
| *ITGB5* | 2,619583 | 9,570096349 | 13,09513933 | 0,000296063 | 0,006401867 |

To investigate the central role of integrins in human TAA pathogenesis, we performed an *in silico* non-standard system biology analysis leveraging public RNA-sequencing databases of aortic MFS-VSMC. First, we considered genes known to be deregulated in primary human MFS-VSMC, shown in red (up-regulated) and blue (down-regulated) in the bottom part of **Supplementary Figure S2**, as deposited by Hansen et al. [29]. Next, we built the protein-protein interaction network comprising high-confidence interactions from well-known public databases. To ensure tissue-specificity, we interrogated the Genotype-Tissue Expression (GTEx) database, which collects updated data on the expression and interactions of genes in specific anatomical niches, relative to human thoracic aorta. This analysis showed that integrins (in yellow) work as hubs of the region of the network responsible for connecting fibrillin-1 to genes deregulated in MFS-VSMC. These data are further depicted in the graph in the inset, which clearly shows that integrins have a high degree of connections (Log_2_ node degree) and a high betweenness centrality value, which measures how often a certain vertex lies on the shortest paths between other vertices, displaying a key role in the connectivity between fibrillin-1 and MFS deregulated genes.

Finally, we validated the protein expression levels of COL1A1 and αV integrin on human thoracic aortic tissues and found a significantly higher expression of both proteins in the media of MFS-TAA and sTAA compared to HC, as shown in **Figure 2**.

**Figure 2.**
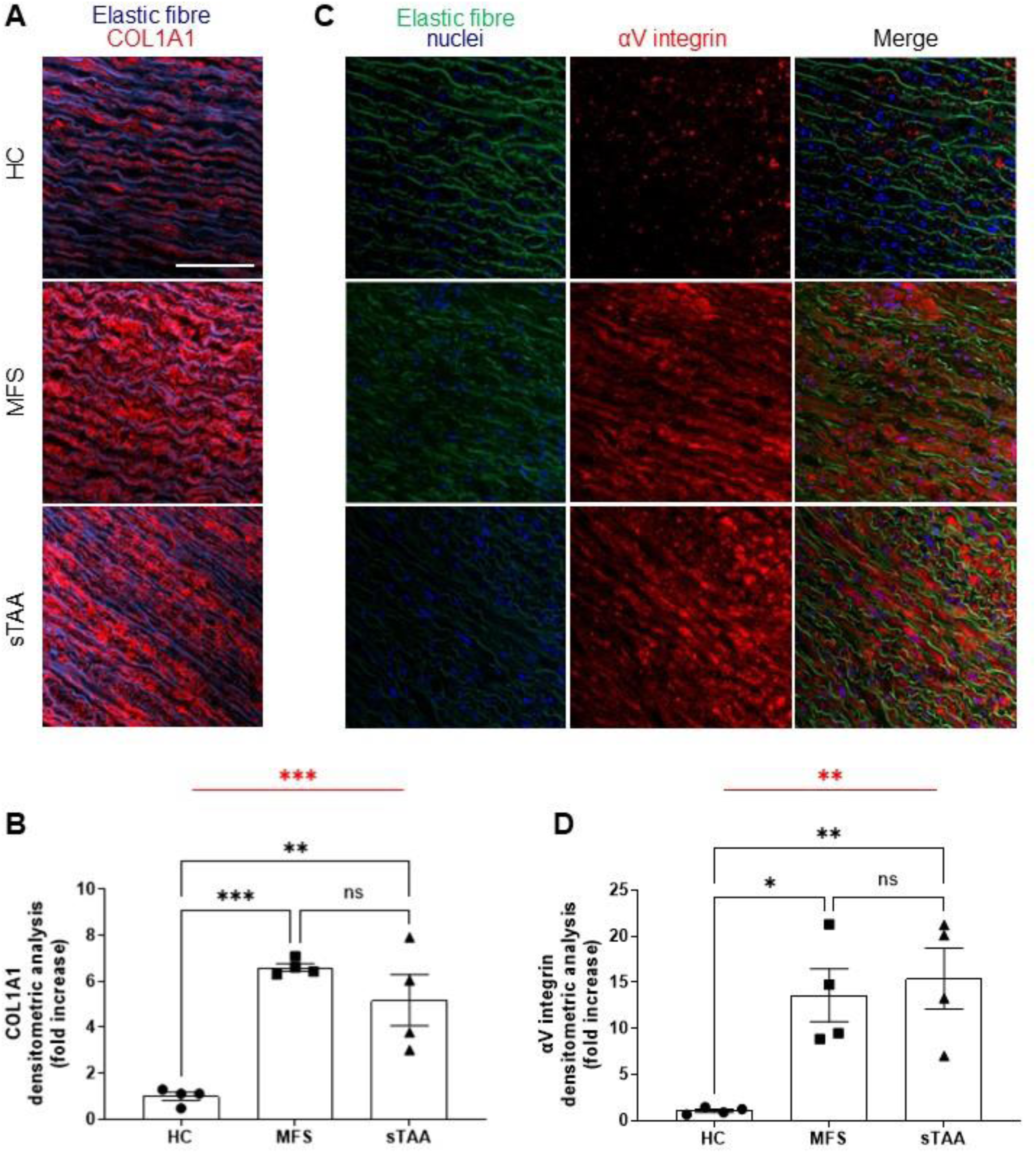
Collagen I and αV integrin are more abundant in MFS-TAA and sTAA than in HC patients. (**A, B**) Representative immunofluorescence images (**A**) and quantification (**B**) of collagen I expression in HC aortas, MFS-TAA and sTAA. (**C, D**) Representative immunofluorescence images (**C**) and quantification (**D**) of αV integrin expression in HC aortas, MFS-TAA and sTAA. Nuclei are stained with DRAQ5 and shown in blue. Scale bar = 100μm. Data are shown as mean±SEM, *n* = 4 per group. One-way ANOVA (in red): \*\**p*<0.01, \*\*\**p*<0.001. Tukey’s *post-hoc* test (in black): \**p*<0.05, \*\**p*<0.01, \*\*\**p*<0.001.

### 3.2 Primary human VSMC from thoracic aortic wall of TAA patients express high levels of αV integrins

We isolated VSMC from MFS and sTAA thoracic aortic wall and compared them to HC-VSMC. We first confirmed the identity of the isolated cells by staining them for the VSMC marker calponin (**Supplementary Figure S3**). Next, we evaluated the expression levels of αV, β5, αVβ3, β1, α5 integrins on the cellular surface, using the ImageStreamX technology, which combines flow cytometry with cell imaging. The percentage of VSMC expressing high levels of αV (**Figure 3A, F**), β5 (**Figure 3B, G**) and αVβ3 (**Figure 3C, H**) was greater in both TAA-VMSC than in HC-VSMC. In contrast, β1 and α5 integrin expression did not vary between groups (**Figure 3D, E, I, J**). Thus, primary VSMC recapitulate the pathological increase in integrin expression previously documented in the transcriptome of aortic tissues from TAA patients.

**Figure 3.**
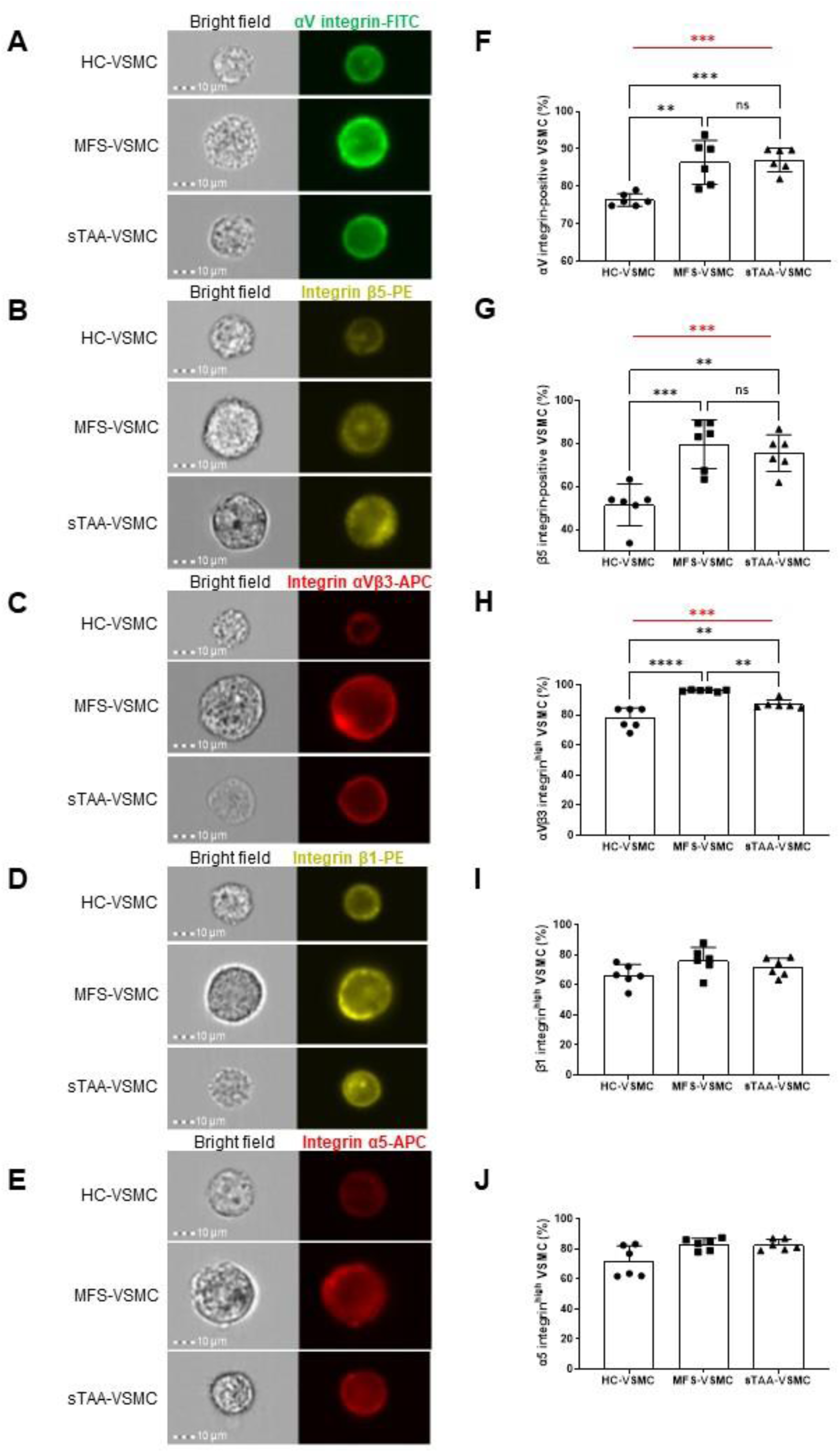
αV, αVβ5, αVβ3, β1 and α5 integrins are more expressed in TAA-VSMC than in HC-VSMC. (**A-E**) Representative ImageStreamX images of HC-, MFS- and sTAA-VSMC in brightfield and stained for αV integrin (**A**), β5 integrin (**B**), αVβ3 integrin (**C**), β1 integrin (**D**) and α5 integrin (**E**). Scale bar = 10μm. (**F-J**) Quantification of the percentage of HC-VSMC, MFS-VSMC, and sTAA-VSMC that express high levels of αV integrin (**F**), β5 integrin (**G**), αVβ3 integrin (**H**), β1 integrin (**I**), and α5 integrin (**J**). Data are expressed as mean±SD, *n* = 6 per group. One-way ANOVA (in red): \**p*<0.05, \*\*\**p=*0.0001. Tukey’s *post-hoc* test (in black): \**p*<0.05, \*\**p*<0.001, \*\*\**p=*0.0001, \*\*\*\**p<*0.0001.

### 3.3 αV integrin inhibition downregulates pro-fibrotic molecular signatures in MFS- and sTAA-VSMC

Our previous data indicate that αV, β3 and β5 integrin subunits are similarly upregulated in MFS- and sTAA-VSMC compared to HC-VSMC. Thus, we explored whether the selective inhibition of αVβ3 and αVβ5 dimers by Cilengitide was effective in reducing pro-fibrotic molecular signatures in cultured VSMC and compared its efficacy with the *pan*-integrin inhibitor GLPG0187.

First, we studied the toxicity of the two compounds by exposing HC-VSMC at multiple doses and measuring cell viability at 72 hours. Based on our data shown in **Supplementary Figure S4A** and **S4B**, we chose the concentration of 0.5 µM for Cilengitide and the concentration of 0.5 ng/mL for GLPG0187 for subsequent *in vitro* experiments. We also validated the safety of the selected doses, either alone or in combination with TGF-β1 (known to play a key role in TAA and here used to mimic pro-fibrotic conditions) on living VSMC. We did not observe significant differences in the number of Annexin V^+^ apoptotic cells (**Supplementary Figure S4C**) or necrotic cells (**Supplementary Figure S4D**) in any of the tested conditions.

As mentioned, αV integrins are known to release the active form of pro-fibrotic TGF-β1 from its latent complex stored in ECM. Therefore, we evaluated the effectiveness of both Cilengitide and GLPG0187 in limiting TGF-β1 release by cultured HC- and MFS-VSMC. **Supplementary Figure S4E** and **S4F** show the significative reduction in the concentration of TGF-β1 released in VSMC supernatant, particularly in MFS patient-derived cells, upon treatment with both αV integrin inhibitors.

Our spatial transcriptomic data indicated that several mediators of pro-fibrotic and mechanotransduction pathways were significantly upregulated in both MFS and sTAA aortic tissues (**Supplementary Figure S5** and **Supplementary Table S3**). Thus, we investigated the effect of integrin inhibitors in limiting the activation of these pathways in primary VSMC isolated from patient, either in basal condition or upon stimulation with human recombinant TGF-β1. **Figure 4** shows representative Western blot images and quantification of activation of both canonical (SMAD2/3) and non-canonical (ERK1/2) TGF-β1 signalling pathways, as well as of downstream mechanotransduction mediators (*i.e.*, RhoA GTPase, ROCK1 and αSMA) in total protein extracts of MFS-VSMC (**Figure 4A-F**) and sTAA-VSMC (**Figure 4G-N**). As expected, treatment with TGF-β1 significantly increased the activation of these pathways in both cell types. Both integrin inhibitors effectively limited the activation of TGF-β1 and mechanotransduction pathway in sTAA-VSMC, whereas Cilengitide was the only drug able to significantly decrease the activation of canonical and non-canonical TGF-β1 signalling pathways in MFS-VSMC. Mechanotransduction was similarly inhibited by both drugs in MFS-VSMC, except for ROCK1 expression levels, which were not modified by Cilengitide. Minor differences were observed in response to both treatments in the absence of TGF-β1.

**Figure 4.**
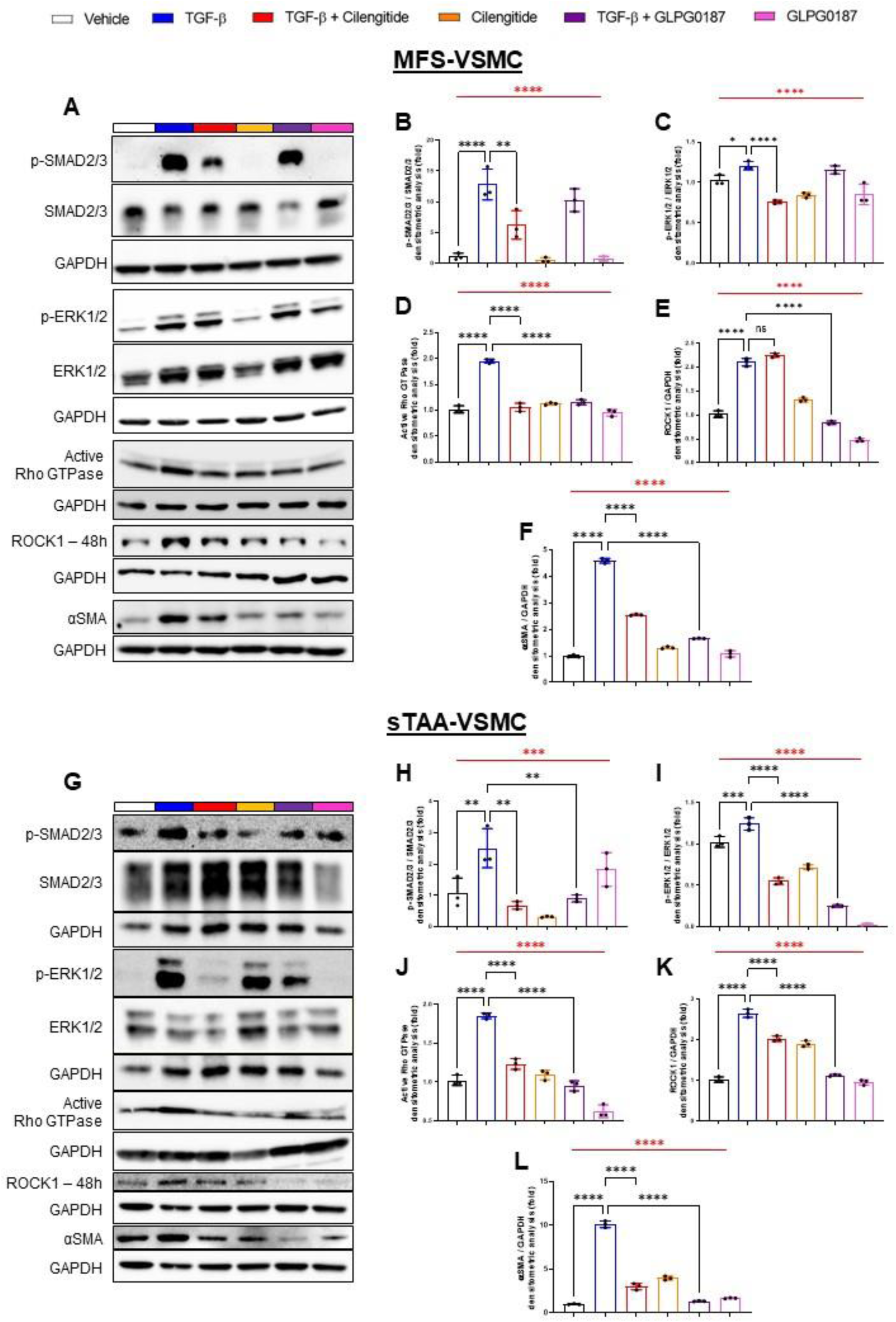
αV integrin inhibition inhibits TGF-β signalling and mechanotransduction pathways in both MFS- and sTAA-VSMC. (**A**) Representative Western Blots for the proteins of interest in MFS-VSMC in response to the indicated treatments. (**B-F**) Densitometric analyses for total:phosphorylated SMAD2/3 ratio (**B**), total:phosphorylated ERK1/2 ratio (**C**), active RhoA GTPase (**D**), ROCK1 (**E**) and αSMA (**F**). (**G**) Representative Western Blots for the proteins of interest in sTAA-VSMC in response to the indicated treatments. (**H-L**) Densitometric analyses for total:phosphorylated SMAD2/3 ratio (**H**), total:phosphorylated ERK1/2 ration (**I**), active RhoA GTPase (**J**), ROCK1 (**K**), and αSMA (**L**). GAPDH has been used to normalize RhoA GTPase, ROCK1 and αSMA expression. Data are shown as mean±SD, *n* = 3 per group. One-way ANOVA (in red): \*\*\*\**p<*0.0001. Tukey’s *post-hoc* test (in black): \**p*<0.05, \*\**p*<0.01, \*\*\**p=*0.0001, \*\*\*\**p*<0.0001.

Thus, in pro-fibrotic conditions αV integrin inhibitors potently limits the activation of both TGF-β1 and mechanotransduction pathways in primary VSMC. Specifically, Cilengitide is more effective than GLG0187 in limiting TGF-β1 activation in both TAA cell types.

### 3.4 αV integrin inhibition downregulates COL1A1 expression levels in MFS- and sTAA-VSMC

Next, we investigated the effect of Cilengitide and GLPG0187 on the expression levels of collagen I (COL1A1), one of the major ECM components responsible for the pathologic fibrotic response in TAA, upregulated in aortic tissues of our spatial transcriptomic data in Figure 1 and known to be under direct transcriptional control of TGF-β1. Since negative feedback loops often result in the downregulation of a receptor in presence of its specific ligand, we evaluated the expression levels of αV integrin by VSMC after treatment with its inhibitors. **Figure 5** shows the expression levels of αV integrin and COL1A1 in MFS- and sTAA-VSMC, as analysed by Western blot (**Figure 5A-F**) and immunofluorescence (**Figure 5G, H**). As expected, the presence of TGF-β1 significantly increased the expression of both αV integrin and COL1A1, in both MFS- and sTAA-VSMC, when compared to vehicle (**Figure 5B-F**). Cilengitide significantly downregulated the expression levels of αV integrin and COL1A1 in all VSMC types, while GLPG0187 was not able to limit COL1A1 expression in sTAA-VSMC.

**Figure 5.**
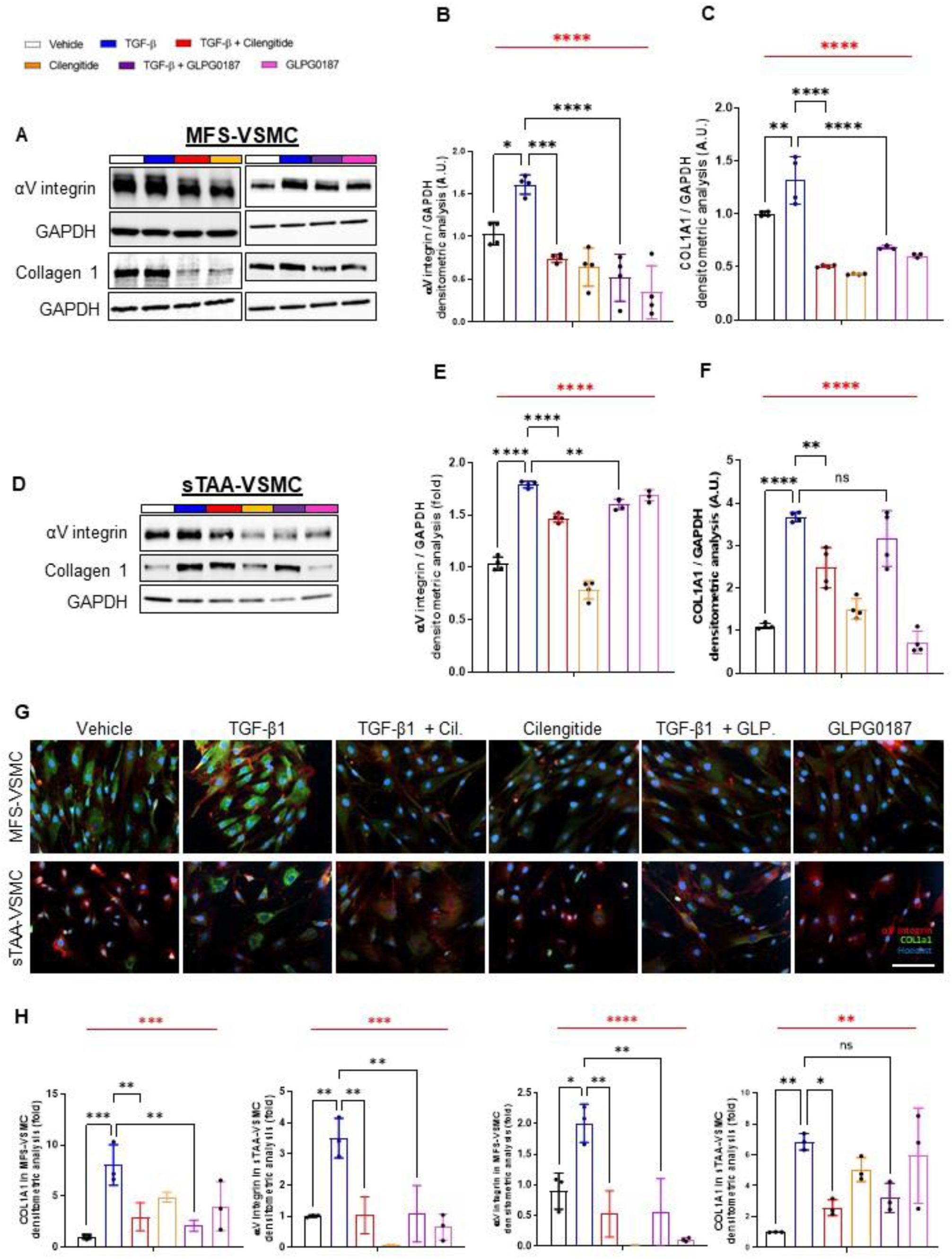
Inhibition of αV integrin downregulates the expression levels of αV integrin and COL1A1 in both MFS- and sTAA-VSMC. (**A-C**) Representative Western blot images (**A**) and corresponding densitometric analysis quantification for αV integrin (**B**) and COL1A1 (**C**) in MFS-VSMC exposed to the indicated treatments. (**D-F**) Representative Western blot images (**D**) and corresponding densitometric analysis quantification for αV integrin (**E**) and COL1A1 (**F**) in sTAA-VSMC exposed to the indicated treatments. GAPDH has been used as protein loading control. Data are shown as mean±SD, *n* = 4 per group. One-way ANOVA (in red): \*\*\*\**p*<0.0001. Tukey’s multiple comparisons *post-hoc* test: \**p*<0.05; \*\**p*<0.01; \*\*\**p*=0.0001; \*\*\*\**p*<0.0001. (**G, H**) Representative images (**G**) and corresponding quantifications (**H**) of immunofluorescence staining for αV integrin and collagen I (COL1A1) in MFS- and sTAA-VSMC exposed to the indicated treatments. Nuclei were stained with Hoechst and are shown in blue. Scale bar = 50μm. Data are shown as mean±SD, *n* = 3 per group. One-way ANOVA (in red): \*\*\**p*=0.0001; \*\*\*\**p*<0.0001. Tukey’s multiple comparisons *post-hoc* test: \**p*<0.05; \*\**p*<0.01; \*\*\**p*=0.0001.

These results were further confirmed in primary MFS patient-specific VSMC, transduced with a lentiviral vector expressing both Enhanced Green Fluorescent Protein (EGFP) and shRNAs specific for ITGAV gene, encoding αV integrin. Efficient VSMC transduction was documented by EGFP expression (**Supplementary Figure S6A**) and reduced levels of ITGAV gene expression (**Supplementary Figure S6B**). Knocked-down MFS-VSMC (αV^KD^) showed reduced expression levels of αV integrin, COL1A1, and TGF-β1 canonical and non-canonical pathway mediators, compared with MFS-VSMC transduced with a scramble shRNA sequence. Treatment with TGF-β1 in αV^KD^ cells resulted in minimal activation of TGF-β1 pathways as well as in a minimal increase of integrin and COL1A1 expression levels.

Thus, αV integrin mediates the activation of TGF-β1 pathways and the production of the pro-fibrotic driver COL1A1. Cilengitide stands as the more effective drug in inhibiting αV integrin and, thereby, the activation of pro-fibrotic pathways in VSMC of all TAA types.

### 3.5 Cilengitide limits thoracic aneurysm progression in both syndromic and non-syndromic TAA murine models

Finally, we tested and compared the therapeutic potential of the two integrin inhibitors in two murine models of TAA, namely Fbn1^C1039G/+^ mice that recapitulate syndromic MFS-TAA, and a model of sTAA, in which lysyl oxidase (LOX) is inhibited by daily administration of β-aminopropionitrile (BAPN) in drinking water for 3 weeks, from the 6^th^ to the 9^th^ week of age, in C57Bl/6 mice [29]. In the syndromic model, we treated both wild-type (WT) and MFS mice 3 times a week, with either Cilengitide or GLP0187, from the 5^th^ to the 16^th^ week of age. In the sTAA model, Cilengitide and GLP0187 were administered 3 times a week, at the same time of BAPN administration.

By 2D-echocardiographic analysis, we monitored the size of two anatomical locations of the ascending aorta, namely the ventriculo-aortic annulus and the Valsalva’s sinus, shown in orange and turquoise in **Figure 6**, respectively. As expected, MFS mice showed statistically higher values for both annulus and Valsalva’s sinus diameters than WT mice at all time points. As shown in **Figure 6B** and **C**, Cilengitide effectively limited aortic dilation at both locations at 18 weeks, whereas GLPG0187 did not show any statistical difference with MFS mice treated with vehicle. BAPN treatment for 3 weeks caused an aortic dilation comparable to the one observed in MFS mice at 16^th^ week of age, confirming the robustness of the model. Also in this case, Cilengitide was the only effective drug in limiting aortic dilation at both locations (**Figure 6E, F**).

**Figure 6.**
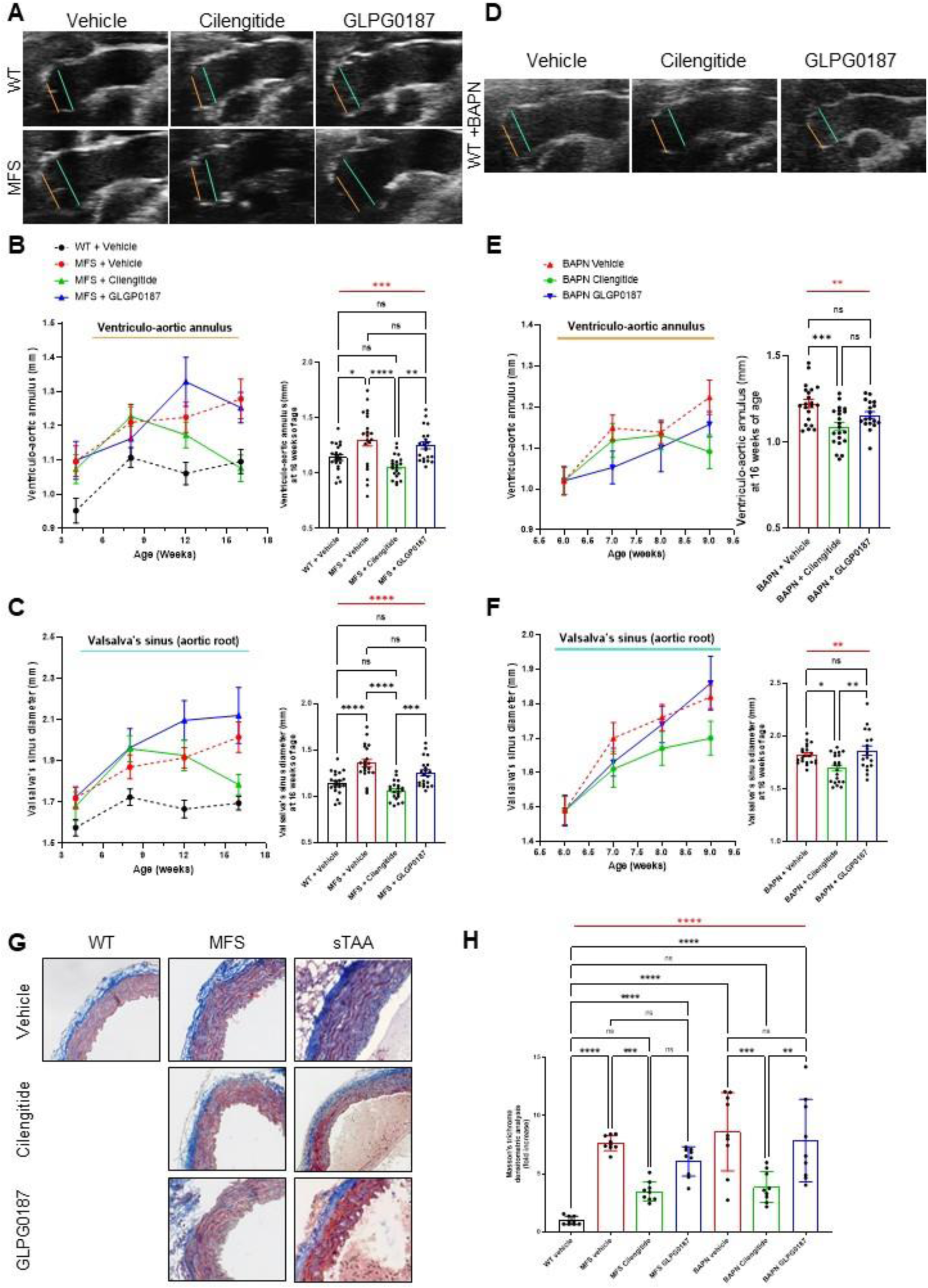
Cilengitide limits thoracic aortic dilatation in both MFS and sTAA murine models. (**A**) Representative 2D-echocardiographic images of ventriculo-aortic annulus (orange line) and Valsalva’s sinus (turquois line) diameters in WT and Fbn1^C1039G/+^ (MFS) mice, treated with vehicle, Cilengitide or GLPG0187. (**B, C**) Quantification of ventriculo-aortic annulus (**B**) and Valsalva’s sinus (**C**) diameters in WT and MFS mice at the indicated time points. Graphs on the right show individual measurements at 11 weeks, in triplicate. *n* = 12 (WT+vehicle); *n* = 13 (MFS+vehicle); *n* = 10 (MFS+Cilengitide); *n* = 11 (MFS+GLPG0187). (**D**) Representative 2D-echocardiographic images of ventriculo-aortic annulus (orange line) and Valsalva’s sinus (turquois line) diameters in mice treated with BAPN in combination with vehicle, Cilengitide or GLPG0187. (**E, F**) Quantification of ventriculo-aortic annulus (**E**) and Valsalva’s sinus (**F**) diameters in BAPN-treated mice in combination with vehicle, Cilengitide or GLPG0187 at the indicated time points. Graphs on the right show individual measurements at 2 weeks of treatment, in triplicate. *n* = 7 (BAPN+vehicle); *n* = 7 (BAPN+Cilengitide); *n* = 6 (BAPN+GLPG0187). All 2d-echocardiographic data are shown as mean±SEM. One-way ANOVA (in red) with Tukey’s *post-hoc* test for multiple comparisons (in black): \**p*<0.05, \*\**p*<0.01, \*\*\**p*<0.001, \*\*\*\**p*<0.0001. (**G, H**) Representative images of Masson’s trichrome staining (**G**) and relative quantification (**H**) of collagen (blue signal) in the *tunica media* of thoracic aortas from WT, MFS or BAPN mice, treated with vehicle, Cilengitide or GLPG0187. Data are shown as mean±SD, *n* = 3, in triplicate. One-way ANOVA (in red) with Tukey’s multiple comparisons *post-hoc* test (in black): \**p*<0.05, \*\**p*<0.01, \*\*\**p*<0.001, \*\*\*\**p*<0.0001.

To evaluate collagen deposition in the aortic wall, we measured the extent of fibrosis in the media layer of thoracic aortas isolated from both MFS and sTAA murine models by Masson’s trichrome staining. This analysis showed reduced collagen deposition in the aorta of both syndromic and non-syndromic TAA mice models after treatment with Cilengitide, but not with GLPG0187, in comparison with vehicle-treated animals, confirming the echocardiography results (**Figure 6G, H**).

Thus, Cilengitide, but not GLPG0187, limits collagen deposition and aortic dilation in both syndromic and sporadic TAA models.

## 4. Discussion

The results described in this manuscript provides fundamental novel insights in our understanding of TAA. First, we provide the first human thoracic aortic spatial transcriptomic set of data from healthy subjects, MFS-TAA and sTAA patients (in both dilated and non-dilated regions). Second, the identified molecular signatures were validated in primary VSMC cultures, obtained from multiple patients. These primary cells are far more representative of the human condition compared to iPSC-derived VSMC that were used in previous studies [23]. Third, we provide proof of concept of the therapeutic potential of Cilengitide, an orphan drug that appears beneficial for both syndromic and non-syndromic TAA. Collectively, these data confirm the central involvement of αVβ3 and αVβ5 integrins in TAA onset/progression and the effectiveness of their selective inhibition in mitigating the disease phenotype in both syndromic and non-syndromic conditions.

The possibility to interfere with integrin activity is becoming a central topic in the management of aortopathies. Recent evidence showed αV integrin overexpression in iPSC-derived aortic VSMC from MFS-TAA patients, proposing the pathway mediated by mTORC as one of the key molecular mechanisms limiting MFS-TAA upon integrin inhibition [23]. Here, we provide evidence for the relevance of additional signalling pathways, such as TGF-β and mechanotransduction, affected by αV integrin inhibition, not only in MFS, but also in sporadic non-syndromic TAA, which pose a higher health and societal burden worldwide.

We and others have contributed to dissect the role of integrins and associated mechanotransduction pathways in other cardiovascular diseases, mainly in hypertensive-derived cardiac fibrosis [30–33]. This led us to apply our findings to TAA [34], for which other evidence pointed to a relevant role of integrins [11, 22, 35]. The recent availability of spatial transcriptomics allowed for a more systematic evaluation of both MFS and sTAA samples. The data presented in this study provide the first characterization of human thoracic aortic specimens and point to a massive expression levels of integrin family members, mechanotransduction, TGF-β signalling pathway and ECM-related genes in both syndromic and sporadic TAA.

Our results suggest that TAA-VSMC undergo a switch from a contractile to a secretory phenotype, which can be effectively reverted by αV integrin inhibition. This issue is debated in the TAA field [36–39] with conflicting results. For example, Pan and colleagues have recently demonstrated that αVβ3 integrin inhibition, mediated by macrophages-derived legumain, leads to Rho GTPase inactivation, which in turn suppresses VSMC phenotypic switch and that this exacerbates thoracic aortic dissection [40]. On the contrary, our results demonstrate that αV integrin inhibition, which limits the switch of patient-derived VSMC into secretory phenotype, downregulates pro-fibrotic molecules and protects from aortic dilation into two different animal models.

Our data align well with recent evidence showing the important involvement of αV integrins in TAA pathology. The “traction model” [12], by which αV integrins binds ECM proteins and activate TGF-β from its latent form stored in the ECM, fits the results of our study and suggests that the main mechanism of action of Cilengitide is mechanotransduction. Our data are consistent with the hypothesis that Cilengitide masks the RGD motif of ECM proteins more effectively than GLPG0187 to αV integrin, with subsequent lowering in TGF-β release from its latent extracellular storage, and reduced VSMC activation. Further structural and docking studies will be necessary to unveil the precise conformation and binding properties of Cilengitide and GLPG0187 to αV integrin and thus to explain the superior efficacy of Cilengitide in preventing aortic dilation in both TAA models. Our data may have implications in other fibrotic pathologies that are still missing of an adequate pharmacological therapy. Indeed, drugs that nowadays reduce fibrosis in organs as lungs, kidneys and heart have so far mainly focused on TGF-β inhibition. However, this approach is fraught by major side effects due to the pleiotropic physiological TGF-β effects [41] and the evidence that TGF-β neutralization may even worsen disease phenotype [41]. In contrast, tackling collateral mechanisms that contribute to the pathology may have significant anti-fibrotic efficacy, with limited side effects [42, 43]. Our results set inhibition of αV integrins as a promising strategy to limit collagen 1 production and thus fibrosis onset/progression. Several molecules have been designed to inhibit integrins by mimicking the RGD amino acid motif [44], mostly in the oncology field. This is also the case for Cilengitide, which has been used in cancer clinical trials [45–49] and, therefore, has been already characterized in terms of pharmacokinetics, pharmacodynamics, efficacy and safety in humans. Its repurposing for the treatment of TAA should therefore be relatively straightforward and easy to bring to the clinics.

## 5. Conclusion

In conclusion, this study paves the way to the use of αV integrin inhibitors for the treatment of TAA. The unique epidemiology of TAA makes Cilengitide a promising target for both a rare syndromic disease (such as MFS-TAA) and a common and widespread condition worldwide, such as non-syndromic sporadic TAA. In this framework, Cilengitide might be the first etiologic drug effectively adopted to limit the fibrosis in the treatment of TAA. Future studies will be necessary to determine, in both TAA cases, if Cilengitide could be offered either alone or in combination with other drugs (*e.g.*, β-blockers, sartans or angiotensin converting enzyme inhibitors), which are routinely used in these patients.

## Author Contributions

Conceptualization, G.L.P.; sampling, G.B., N.S.U., J.T., A.P.; experiment, G.L.P., S.R., S.B., N.F., M.C., S.V., F.R., F.C., L.L., M.C.S., L.E., M.V., L.P., R.S., V.M.; writing—original draft preparation, G.L.P., S.R., S.B, L.E., L.P.; writing—review and editing, G.L.P., S.Z., G.P., P.P.; supervision, G.L.P.; funding acquisition, G.L.P., S.Z., G.P., P.P., L.E. All authors have read and agreed to the published version of the manuscript.

## Acknowledgement

We would like to thank the genomic facility at IRCCS Humanitas for the technical support with spatial transcriptomic experiments.

## Funding

This research was supported by the Italian Ministry of Health funds (Ricerca Finalizzata GR-2021-12371950 and RC2022-4.03 ID: 2771963), Fondazione Gigi e Pupa Ferrari ONLUS (FPF-14) and by Italian Ministry of Research (PRIN 2022, No. 2022Y849WY).

## Informed Consent Statement

Informed consent was obtained from all subjects involved in the study. Written informed consent has been obtained from the patient(s) to publish this paper.

## Conflicts of Interest

The authors declare that they have no competing interests.

## Notes

### Competing Interest Statement

The authors have declared no competing interest.

### Author Declarations

All patients signed and gave written informed consent for tissue collection for research purposes and the procedures were approved by Ethics Committee either of Centro Cardiologico Monzino IRCCS (RE3001-CCM 1462) and ASST FBF-Sacco (Prot. N. 39138/2016, Milan, Italy).

